# Prevalence and experience of fatigue in survivors of critical illness: A mixed-methods systematic review

**DOI:** 10.1101/2020.06.30.20138248

**Authors:** Suzanne Bench, Louise Stayt, Akshay Shah, Paula Dhiman, Wladyslawa Czuber-Dochan

## Abstract

**Objective:** To investigate the prevalence, experience and management of fatigue in adult survivors of critical illness.

**Data sources:** An electronic search was performed on seven databases: CINAHL, Medline, EMBASE, PsycINFO, Emcare, British Nursing Index, and Web of Science.

**Study selection and data extraction:** Seventy-six peer-reviewed, English language studies that investigated fatigue or vitality in adult patients admitted to an Intensive Care Unit (ICU) and followed up for any period were included. Extracted data were split into three datasets for analysis: data from vitality domain of SF-36 (dataset A); other quantitative data (dataset B); and qualitative data (dataset C). Methodological quality was assessed using the critical appraisal skills programme (CASP) tools.

**Data synthesis:** We adopted a segregated approach to mixed-methods synthesis. We merged all datasets, attributing all results to one of four qualitative themes: prevalence and severity; contributing factors; impacts on quality of life; assessment and management.

**Results:** The prevalence of fatigue ranged from 13.8 to 80.9%; fatigue severity reached its nadir at approximately one-month post-ICU discharge, improved over time but seldom reached reference population scores. Various biological, disease-related and psychological factors associated with fatigue were identified including age, poor pre-morbid status and sleep disturbance. No critical illness specific tool to assess fatigue in ICU survivors was identified. A paucity of evidence-based interventions for management of fatigue in ICU survivors were identified despite fatigue negatively impacting on survivors’ quality of life.

**Conclusions:** This mixed method review shows that fatigue is highly prevalent in ICU survivors, negatively impacting their recovery. To date, no ICU specific fatigue assessment tool or targeted intervention has been designed to manage this symptom. Our review has identified factors, which may increase or mitigate against fatigue, along with potential management strategies, which should be used to inform future research and practice.

## INTRODUCTION

Approximately 130,000 patients survive an intensive care unit (ICU) admission in the United Kingdom (UK) every year. (1) Survivors commonly report long-lasting physical, cognitive and psychosocial problems impacting their quality of life, a combination often termed post-intensive care syndrome (PICS). (2,3) PICS can also impact on the family members of ICU survivors. (4,5) A cardinal symptom of PICS is fatigue, (6) which is defined as an overwhelming, sustained sense of exhaustion, typically unrelieved by sleep, with decreased capacity for physical and mental work at a usual level. (7,8)

Recent data suggest that fatigue is an important but under-recognised problem in ICU survivors. (9-12) In a qualitative study, patients’ ranked fatigue as one of their three most important outcomes. (13) International advisory panels have called for research into the prevalence, severity, underlying mechanisms and management of fatigue in ICU survivors. (14) Moreover, although the long-term consequences of Covid-19 are unknown, preliminary reports suggest that many survivors will experience a post viral chronic fatigue syndrome. (15)

Previous work has evaluated overall health-related quality of life (HRQoL) following ICU and report some data on critical illness fatigue. (6,16) In addition, a narrative review on the assessment and management of fatigue in ICU has been published. (17) The aim of this systematic review was to identify the prevalence, experience, risk factors and management of fatigue in adult ICU survivors.

## MATERIALS AND METHODS

This systematic review was conducted according to a study protocol registered on PROSPERO (CRD42018091992). We report our findings in accordance with the preferred reporting items for systematic reviews and meta-analysis (PRISMA) statement. (18) We undertook a mixed-methods approach combining studies from different research methodologies and undertook a segregated approach to mixed-methods synthesis. (19)

### Eligibility criteria

We considered primary research of any methodology published in English. Studies that investigated fatigue in patients admitted to an adult ICU and followed up for any period were included. We excluded critically ill neonates and children, studies that focused on fatigue secondary to a solitary pathological process (e.g. brain injury) and those on a different but parallel topic (e.g. sleepiness). Studies reporting data collected whilst the patient was still in the ICU were also excluded. Due to the extensive number of studies reporting Medical Outcomes Study 36-item Short Form Health Survey (SF-36) data, only papers published after 2000, which mentioned ICU in the abstract and reported specific vitality data as a general measure of fatigue were included.

### Study identification and selection

We searched seven databases: CINAHL®, MEDLINE®, EMBASE®, PsycINFO®, OVID® Emcare, British Nursing Index and the Web of Science™ from 01 Jan 1946 until 28 Feb 2018 (updated 14.05.2020). The full search strategy is available as supplementary digital content: 1. We also contacted known experts and searched professional websites using the terms fatigue and vitality. We performed forward and backward citation searches on all studies that met the inclusion criteria.

A single reviewer (S.B.) screened all titles and abstracts to assess eligibility for full-text review. The full text of selected studies was independently reviewed by at least two authors to ensure they met eligibility criteria. Any discrepancies were resolved through discussion and consensus.

### Data extraction and quality assessment

Data were extracted by the authors on pre-piloted data extraction forms. Methodological quality was assessed using the critical appraisal skills programme (CASP) tools. (20) No study was excluded on the basis of its methodological quality (supplementary digital content: 2), but results were used to assign each study a grade (green, amber, red) based on the quality and strength of the evidence (Table 1). Consensus agreements by the whole team were used to determine final decisions.

**Table 1:**
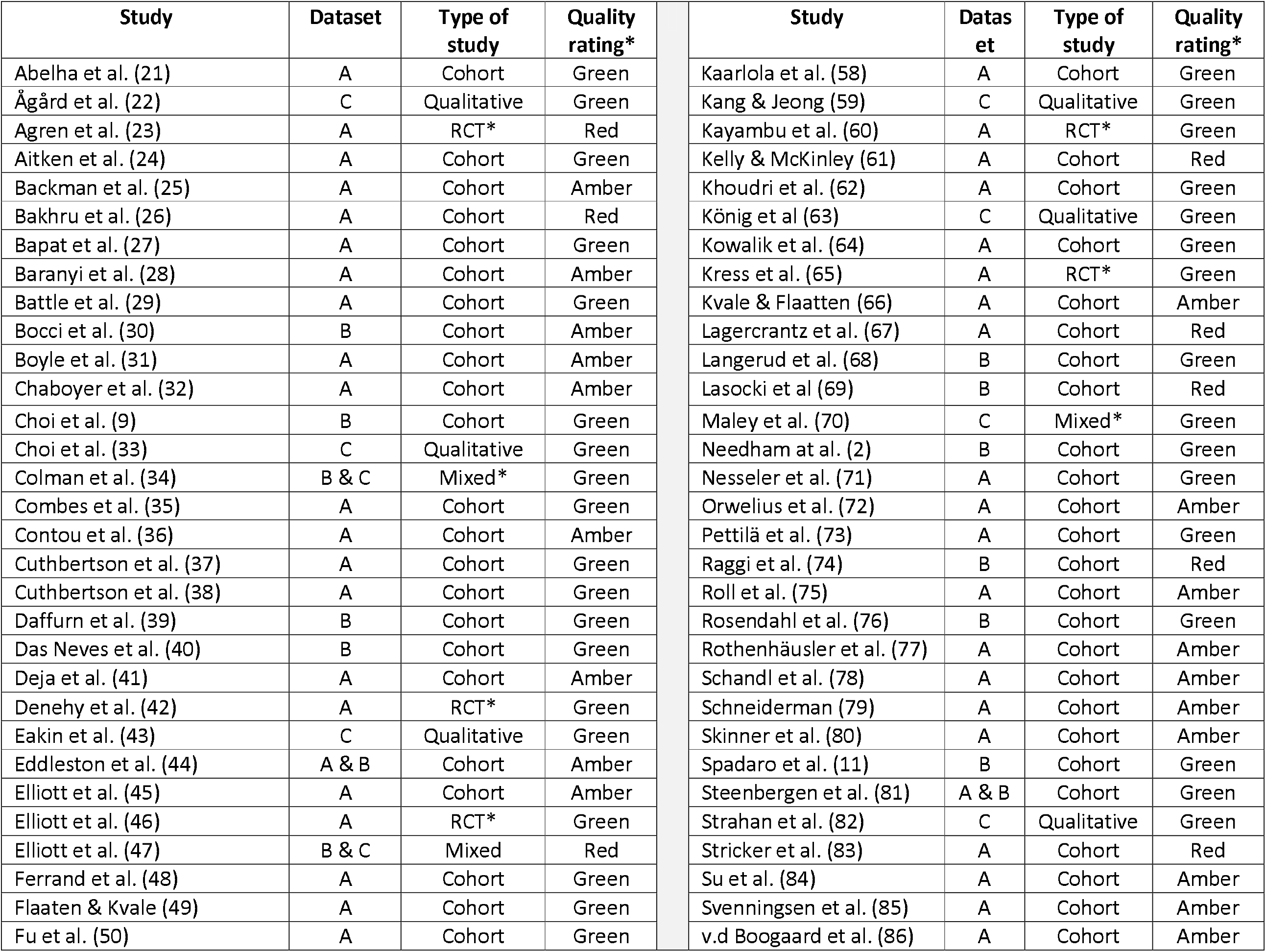
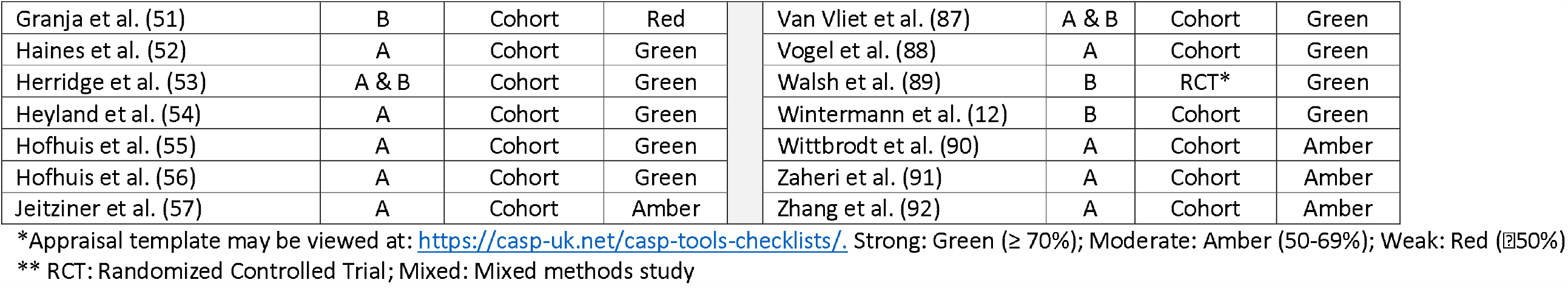
Included studies and their methodological quality rating

### Data synthesis and analysis

We adopted a segregated approach to mixed-methods synthesis. (19) Extracted data were split into three datasets for analysis: data from vitality domain of SF-36 (dataset A); other quantitative data (dataset B); and qualitative data (dataset C).

Mean SF-36 vitality domain scores (dataset A), standard deviation and sample size were extracted for each reported time point. Mean vitality scores were combined to produce a weighted mean score. Indication of ICU admission type was categorized as: unselected general cohort; sepsis; or surgery. The weighted mean vitality score, standard deviation and 95% confidence interval (CI) were presented for each study design. Studies presenting median SF-36 vitality score were not included in this analysis. We used STATA for analyses (Version 15; StataCorp, College Station, Texas, USA). Pooling of results from other quantitative data (dataset B) was not possible due to the heterogeneity of assessment tools used to measure fatigue thus results are presented narratively.

Qualitative data (dataset C) were thematically analyzed. (93) In a final step, we merged all datasets, attributing all results to one of the qualitative themes. Results are reported under these themes: prevalence and severity; contributing factors; impacts on quality of life; assessment and management.

## RESULTS

### Study selection and characteristics

Seventy-six studies were considered eligible (Figure 1).

**Figure 1:**
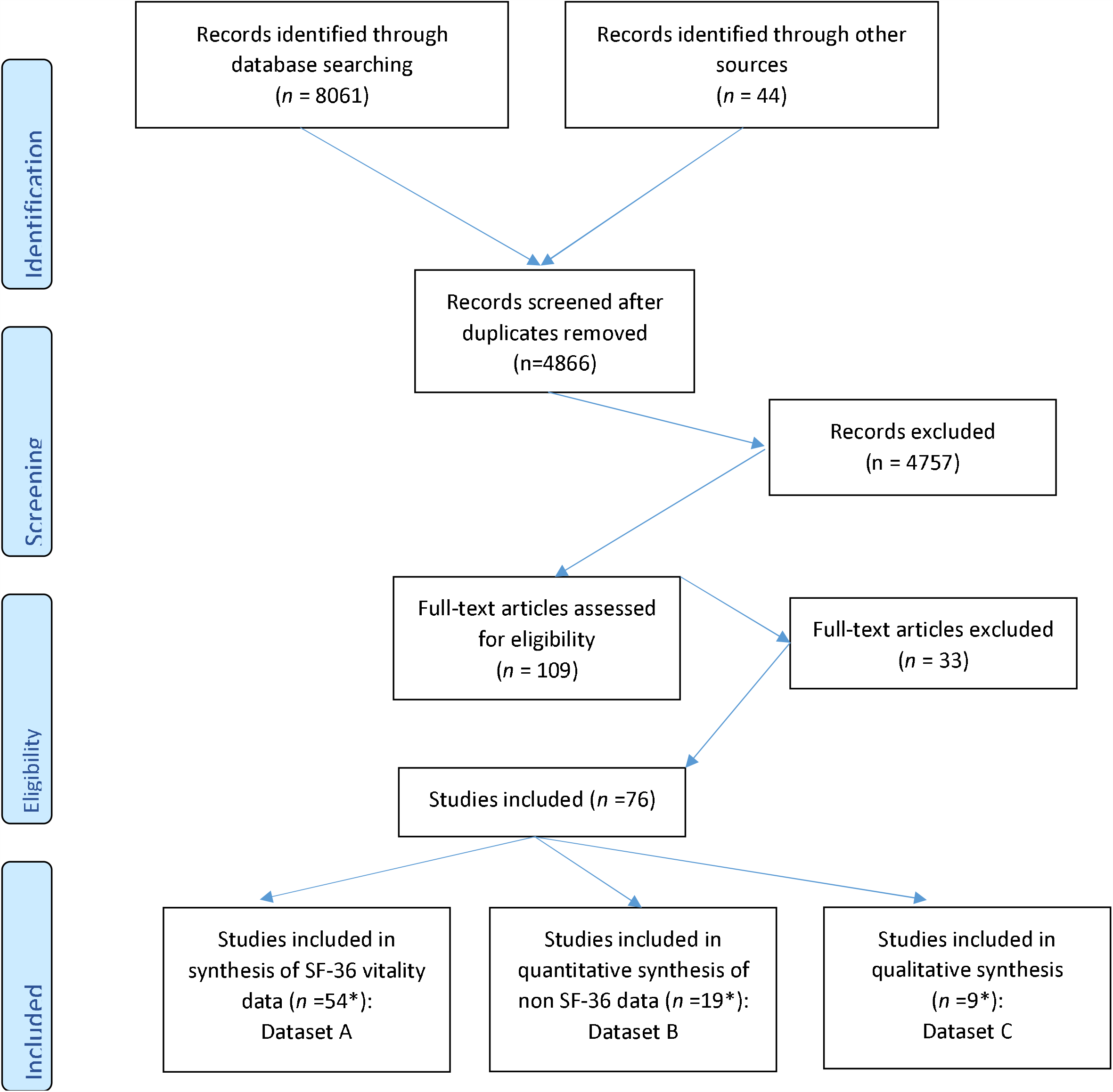
Prisma Diagram. *Some studies included data relevant to more than one synthesis approach, thus numbers from each dataset do not equate to total studies included

Sixty-one included studies were observational, six were randomized controlled trials (RCTs), six were qualitative and three were mixed methods studies (Table 1). Forty-four studies were conducted in Europe, 13 in Australasia, seven in North America, and eight in other parts of the world (Argentina, China, Iran, Morocco, South Africa, South Korea). Majority of studies (*n*=53 (73%)) were single center investigating a general/unselected ICU patient cohort (*n* =45 (62%)). Only one of the qualitative studies focused specifically on fatigue. (34) All others evaluated fatigue as part of a wider focus on HRQoL after critical illness. Two qualitative studies also report data from the perspective of relatives. (22,33) Full details of included studies can be found as supplementary digital content: 3a and 3b.

Fifty-four studies provide SF-36 vitality data and 19 report other quantitative data using a range of fatigue assessment tools (supplementary digital content: 4). Follow up assessment was most commonly evaluated 6-12 months after ICU or hospital discharge Nine studies evaluated outcomes at two or more years following hospital discharge. Only two studies collected pre-ICU/hospital admission vitality data and eight studies collected vitality data at the point of ICU discharge (Supplementary file 5).

### Quality of the evidence

Studies were generally of adequate quality; defined by a subjective rating of amber or green (supplementary digital content: 2). Follow up rates for SF-36 studies exceeded 70% in 25 (52%) studies, with a median (IQR) response rate of 71.5% (48.7 – 82.3). Response rates in dataset B ranged from 35% (40) up to 100%. (34, 39) Response rates were higher in studies that used face-to-face assessment or a combination of methods (supplementary digital content: 5). However, few observational studies adequately identified and considered all confounding factors. This was attributable to vitality or fatigue often being a secondary outcome measure. Several qualitative studies provided insufficient data to allow a full judgement of quality. Regardless of methodological quality ratings, all data were treated equally during analysis.

### Synthesis of results

#### Prevalence and severity

The reported prevalence of fatigue ranged from 13.8% at one year to 80.9% four months post-ICU discharge. (9,32,51,68,70) Vitality scores reached a nadir at one month following ICU discharge and slowly improved over time (Table 2 & Figure 2A) but remained worse than the reference population in most studies until follow-up was complete. Vitality scores obtained from RCT data were lower than those from cohort studies (Figures 2A and 2B).

**Table 2:** SF-36 Vitality scores of included studies over time

| Study design<br>(*n* = number<br>of studies) | Baseline | 1 month | 3 months | 6 months | 9 months | 12 months | 24 months | 60 months |
| --- | --- | --- | --- | --- | --- | --- | --- | --- |
|  | (n = number of study participants providing vitality data) |  |  |  |  |  |  |  |
| Cohort<br>(*n* = 38) | 49.71 (25.75)<br>[48.44 to<br>50.98]<br>(*n* = 1586) | 46.18 (22.80)<br>[44.48 to<br>47.88]<br>(*n* = 690) | 53.56 (22.72)<br>[52.36 to<br>54.76]<br>(*n* = 1370) | 55.40 (24.05)<br>(54.39 to<br>56.41)<br>(*n* = 2194) | UA | 53.78 (24.07)<br>(52.83 to<br>54.73)<br>(*n* = 2464) | 55.69 (22.13)<br>(54.61 to<br>56.77)<br>(*n* = 1610) | 57.02 (22.29)<br>(54.79 to<br>59.25)<br>(*n* = 387) |
| RCT<br>(*n* = 5) | 38.91 (12.99)<br>(36.43 to<br>41.39)<br>(*n* = 108) | UA | 42.80 (12.02)<br>(40.44 to<br>45.16)<br>(*n* = 102) | 43.45 (13.92)<br>(41.17 to<br>45.73)<br>(*n* = 145) | UA | 45.65 (12.91)<br>(42.70 to<br>48.38)<br>(*n* = 82) | UA | UA |
| Cross-<br>sectional<br>(*n* = 8) | UA | 56.44 (32.3)<br>(52.92 to<br>59.97)<br>(*n* = 325) | 50 (18.5)<br>(46.96 to<br>53.04)<br>(*n* = 145) | UA | UA | 54.66 (16.1)<br>(52.83 to<br>56.49)<br>(*n* = 299) | UA | UA |
| Case-control<br>(*n* = 2) | UA | UA | UA | UA | UA | 71.63 (18.86)<br>(67.29 to<br>75.97)<br>(*n* = 75) | UA | UA |
| Before-and-<br>after<br>(*n*=1) | UA | UA | UA | UA | 10.08<br>(*n* = 19) | UA | UA | UA |

Values denote Mean (SD) [95% Confidence Interval], UA = Unavailable sufficient data on mean or SD.
RCT, Randomised Controlled Trial

**Figure 2A:**
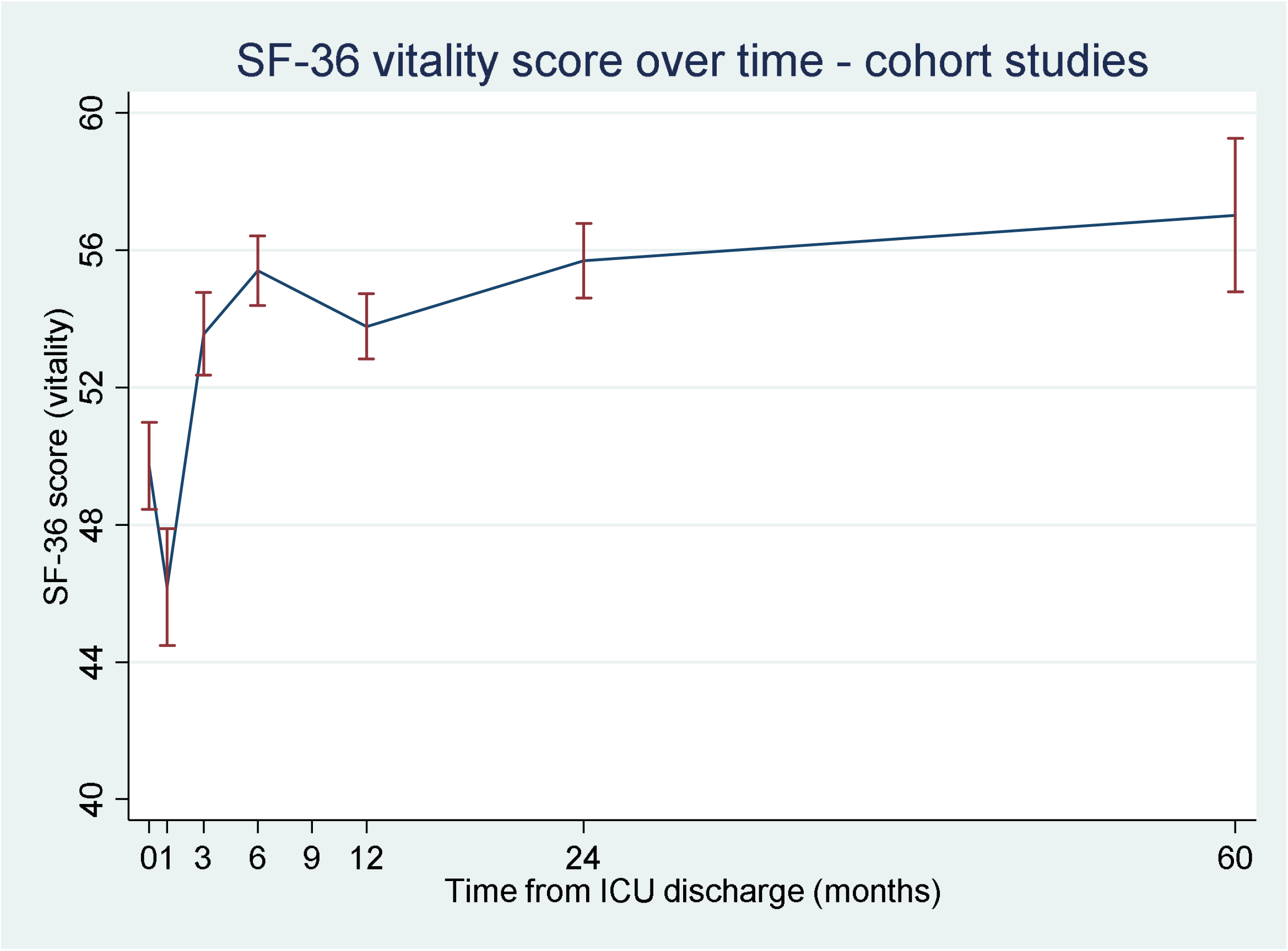
Mean (95%CI) SF-36 vitality scores over time for data from observational cohort studies

**Figure 2B:**
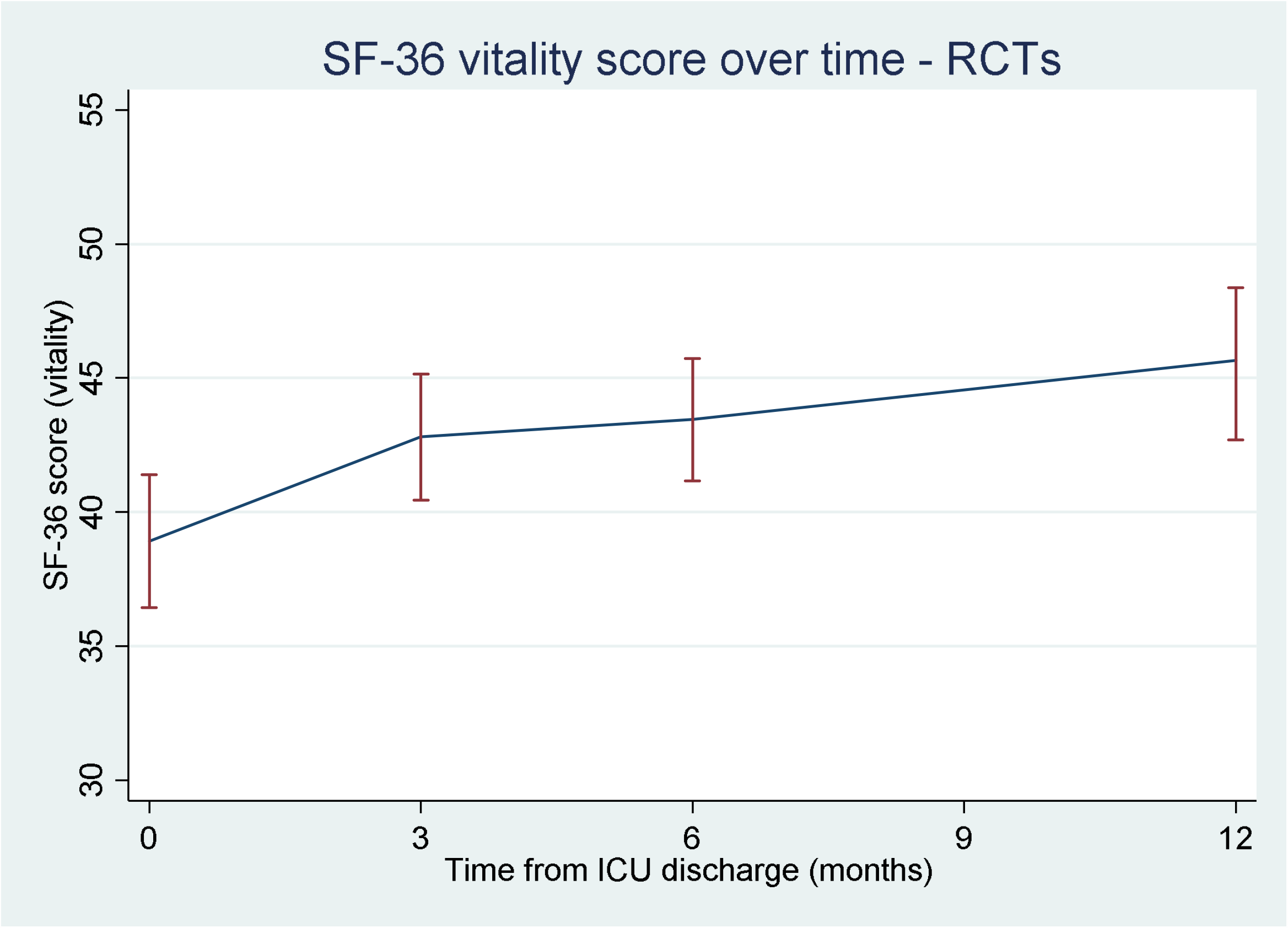
Mean (95%CI) SF-36 vitality scores of data obtained from randomized controlled trials

Qualitative findings support fatigue as a commonly experienced symptom post ICU discharge, with people describing it as a complex symptom rather than simple muscle weakness. (59) Fatigue was particularly prevalent in the early period after ICU discharge, (22,34,82) and for many people, fatigue symptoms and vitality improved over time. (43,63) Fatigue was generally viewed as an expected and integral part of recovery: “I *just think of it as getting over what I’ve been through*”. (34) However, recovery from fatigue can take time and survivors were surprised by this: “…*I am similarly stunned at the time it’s taken to get to the point where I am at”*. (47)

#### Contributing factors

A range of factors are reported to be associated with fatigue following ICU discharge and these are summarized in Table 3. However, these were not consistently observed across all studies.

**Table 3:** Factors associated with fatigue in ICU survivors

| Negative impact | Positive impact |
| --- | --- |
| Patient / demographic<br>• Female sex ($n = 3$) (21,44,50)<br>• Age – both increasing age (12,48,50,57,91) and young age, especially in males (21) ($n = 6$)<br>• Poor pre-morbid vitality / quality of life scores ($n = 3$) (12,37,51, 74)<br>• High pre-existing co-morbidity ($n = 2$) (12, 72) | • Psychoeducational ($n = 1$) (23)<br>• Increased 6-minute walking distance ($n = 1$) (24)<br>• ICU diaries ($n = 1$) (25)<br>• Mild therapeutic hypothermia (following out of hospital cardiac arrest) ($n = 1$) (64) |
| Admission/ICU-related<br>• High ICU admission illness severity scores ($n = 3$) (48,50,62)<br>• Multiple organ dysfunction ($n = 1$) (73)<br>• Severe sepsis/septic shock ($n = 2$) (29,92)<br>• Prolonged ventilation ($n = 2$) (48,91)<br>• ICU length of stay ($n = 2$) (27,50)<br>• Hydroxyethyl starch fluid resuscitation ($n = 1$) (90)<br>• Traumatic brain injury ($n = 1$) (50)<br>• Cognitive impairment ($n = 1$) (77)<br>• Muscle weakness ($n = 4$) (9,30,53,69)<br>• Iron deficiency ($n = 1$) (69) |  |
| Psychological / constitutional<br>• Pain ($n = 5$) (9,30,31,39,68)<br>• Sleep disturbance ($n = 6$) (9,30,39,44,47)<br>• Depression and/or anxiety ($n = 4$) (12,30,68,74)<br>• PTSD or PTSS ($n = 2$) (12,68)<br>• Breathlessness / dyspnoea ($n = 1$) (11)<br>• Weight loss ($n = 1$) (53) |  |
| Social<br>• Lack of social support ($n = 1$) (12)<br>• Discharged home following ICU ($n = 1$) (9)<br>• Unable to return to employment ($n = 1$) (53) |  |

### Impact on quality of life

Fatigue is reported to have a profound impact on cognitive, physical and social dimensions of an individual’s functioning. (34) Fatigue is also associated with a significantly lower Barthel Index at discharge (12) and is a commonly cited cause of reduced physical function (61) as described by one person who said: *“I can’t walk very far. I’ve* just got no energy”. (47) This in turn affects people’s independence with regards to their personal care; one participant said: *‘‘… somebody has to take me for a shower and that exhausts me”*. (82) Fatigue also impacts on wider activities, highlighted in the following quote: *“I can only do one thing a day. If I had two appointments, I couldn’t make it because I would be exhausted even before I finished the first one”*. (59) Long-term iron deficiency was also reported to impact fatigue preventing a return to pre-ICU admission daily activities. (81)

Fatigue is associated with a greater risk of being diagnosed with depression. (13) Fatigue can mean survivors lose their identity and their self-worth because they are unable to look after themselves or to perform their normal social roles, such as being a parent or partner. (22,34) Fatigue affects both employed and retired participants’ ability to return to their previous level of activity. (51) Fatigue also has a financial impact: *“I’d lost the business, … we were in debt to the bank*… *We had no money coming in, we couldn’t pay the mortgage… Just all those money worries”*. (34) Being unable to work also impacted on people’s status within the family and made them feel a burden. (59)

Survivors often had little energy for social activities such as interaction with friends and family. (22) The social impact was made worse by what was described as ‘cognitive fatigue’, leaving people with difficulties with concentration, memory and thought processing: *“I would think, oh, I wish this was over. I want to go home and have a sleep…. things like laughing and being humorous…that’s not really important when you’re trying to do the basics of having a conversation”*. (34)

### Assessment and management of fatigue

In addition to the SF-36, 11 tools were used, either in their original form or as a modified version, to measure the presence, severity or impact of fatigue (supplementary digital content: 4). Tools varied in length from 40-items (Fatigue Impact Scale (FIS)) to 20- or 18-items (Multidimensional Fatigue Inventory-20 (MFI-20) and Lee Fatigue Scale (LFS) respectively), down to 13, 9 or 8 items (Functional Assessment of Chronic Illness Therapy for Fatigue (FACIT-F), Fatigue Severity Scale-9 (FSS-9), Checklist individual strength-fatigue (CIF-F)), with some being just a single item (e.g. Visual Analogue Scale (VAS)).

Some tools were designed to solely measure fatigue, while other tools had a sub-section or one question designated to assess fatigue, or related constructs. Different scales provided different information on fatigue. This ranged from a simple Yes/No answer on tools such as the Symptom Assessment Tool (SAT), or a rating of severity using, for example, a VAS numerical scale to more discreet severity scores for different fatigue domains such as general fatigue, physical fatigue, mental fatigue, reduced motivation, and reduced activity (MFI) or cognitive, physical and psychosocial impact of fatigue (FIS).

Causes of fatigue were only assessed in two studies using the FSS-9 (48,74) and only three studies used one of two tools (FIS and FSS-9) to measure the impact of fatigue. (34,47,74) None of the tools were developed with ICU survivors and only two, FACIT-F scale and MFI-20, were validated in ICU patients. Spadaro et al. report that the reliability and construct validity data they collected suggest that the FACIT-F scale grasps the negative aspects of fatigue better than the vitality dimension of SF-36, (11) whilst Wintermann et al. report the MFI-20 to have a Cronbach’s *α* of 0.91. (12)

People reported a range of strategies to mitigate and manage their fatigue. As well as trying to eat well and taking regular naps to avoid feeling *“wiped out”* (22,34,43), exercise was seen as beneficial as *“any tiredness I had after that [exercise] I felt was a natural tiredness, not just a tiredness from being unwell”*. (34) This included trying to exercise the brain by doing things like puzzles, although the ability to do this was limited by the fatigue itself: *“When I play it [Sudoku] and the time it takes for me to do it is all related to the fatigue factor and the concentration factor so if I am fatigued it takes forever to do it and I just have to put it down”.* (47)

Pacing activities and prioritizing were reported as strategies to manage fatigue. (34,43,59) Planning ahead and being organized helped people to continue with their daily activities: *“I do have to write on the calendar… I had the whole week planned… and I had to write it all down to make sure I knew exactly what I was doing”.* (43)

Finally, education and information about fatigue, its impacts and how it can be managed was considered important, but difficult to obtain: *“Nobody forewarned us about anything…. Even if a doctor sat you down and said to you ‘you can expect to be very tired for the next two years. You’re going to get fatigue… Expect this”, whilst another said “The fatigue part of it has never been broached. Never”.* (34)

## DISCUSSION

### Key findings

In this most comprehensive review to date, we have demonstrated that: (i) fatigue is common in ICU survivors with a prevalence ranging from 13.8 to 80.9%; (ii) fatigue severity reaches its nadir at approximately one month post-ICU discharge, improves over time but seldom reaches reference population scores; (iii) there is no critical illness specific tool to assess fatigue in ICU survivors; and (iv) there is a paucity of evidence-based interventions for management of fatigue in ICU survivors despite fatigue having a profound negative impact on ICU survivors’ quality of life.

Our findings support those from studies with other long-term conditions, including cancer, (94) inflammatory bowel disease, (95) and chronic kidney disease (96), supporting fatigue as a commonly experienced symptom of ill health.

### Strengths and limitations

Employing a mixed-methods approach enabled us to produce a comprehensive review of all available evidence, with estimates that could be useful to inform power calculations for future studies (Table 2). Our review also identifies factors, which may increase or mitigate against fatigue (Table 3) which future researchers might find useful.

Our review has limitations. Meta-analysis of the vitality data was not possible due to the degree of heterogeneity. Additionally, differences in study design, patient populations, fatigue measurement tools, follow-up time points and response rates of the studies included in our review make it difficult to provide one overall conclusion.

### Recommendations for future practice and research

Fatigue is multifaceted, with physical and mental components and multifactorial, related to a variety of modifiable and non-modifiable factors. Further study, particularly qualitative, is needed to better understand patient fatigue and its impact on quality of life.

The variety of scales used to assess fatigue make it difficult to compare severity, types and impact between studies and across patient populations. We recommend the development of a critical illness specific fatigue assessment tool. Most tools used to assess fatigue have been developed for and validated in other population groups, for example, cancer, chronic fatigue, inflammatory bowel disease and stroke. (97-100) Two fatigue assessment tools have been validated in critical care, (11,22) however, none have been developed with ICU survivors.

The variable prevalence of fatigue could be due to the different tools used for assessment and the different time points for measuring outcomes. Fatigue severity reaches a nadir at one-month post-ICU discharge and demonstrates the greatest improvement in the first year after discharge (Fig. 2A). Therefore, interventions to treat fatigue in critical care survivors might be most effective in this time period.

To address its multidimensional nature, the management of fatigue requires development of a complex intervention. Findings of our review and those with other population groups suggest a tailored, multifaceted approach with recommendations for nutrition, exercise, pacing activities and education/information. (101-105) Outside of critical care, other non-pharmacological and pharmacological interventions including alternative therapies (106) and the use of iron, modafinil and doxepin, have also been tested, with the latter two proving effective in patients with Parkinson’s disease. (107)

The estimates in this review can be used to inform power calculations for future long-term trials, which should include collection of pre-ICU fatigue/vitality data for comparison where possible. Conducting long-term outcome research in ICU survivors is challenging, however, more than half of included studies in our review had follow up rates of greater than 70%.

Finally, the impact of critical illness on family members’ fatigue remains an unexplored area and is a strong recommendation for future research. Despite Choi et al. reporting that fatigue is also experienced by family members, (33) our original search failed to uncover enough data to review further.

## CONCLUSION

This mixed method review shows that fatigue is highly prevalent in ICU survivors, negatively impacting their recovery. To date, no ICU specific fatigue assessment tool or targeted intervention has been designed to manage this symptom. Our review has identified factors, which may increase or mitigate against fatigue, along with potential management strategies, which should be used to inform future research and practice.

## Data Availability

Data are available on request to the corresponding author

## Acknowledgements

We acknowledge the support of Dr Micol Artom, who assisted with data collection and screening.

## SUPPLEMENTARY FILES

Supplementary Digital Content 1: Search strategy and results

Supplementary Digital Content 2: Quality appraisal

Supplementary Digital Content 3a: Study characteristics

Supplementary Digital Content 3b: Details of qualitative studies

Supplementary Digital Content 4: Fatigue assessment tools

Supplementary Digital Content 5: Follow up and response rates

